# An Environment-Wide Study of Adult Cognitive Performance in the 23andMe Cohort

**DOI:** 10.1101/19009076

**Authors:** Yunru Huang, Teresa Filshtein, the 23andMe Research Team, Robert Gentleman, Stella Aslibekyan

**Author notes:** **Materials and Correspondence**: Stella Aslibekyan.

## Abstract

**BACKGROUND:** With the emergence of web-based data collection methods, large digital health cohorts offer the opportunity to conduct behavioral and epidemiologic research at an unprecedented scale. The size and breadth of such data sets enable discovery of novel associations across the phenotypic spectrum.

**METHODS:** We deployed the digital symbol substitution test (DSST) online to consented 23andMe research participants 50-85 years of age. We tested cross-sectional associations between DSST performance and 824 phenotypes using linear regression models adjusted for age, sex, age*sex interaction, device, time of cohort entry, and ancestry, separately among discovery (n=144,786) and replication (n=93,428) samples; additional analyses further adjusted for education. We post-stratified association estimates on age, sex, and education to adjust for discrepancies across subsamples. Leveraging the rich genetic and phenotypic data available at 23andMe, we also estimated genetic and environmental correlations between DSST and its top correlates using linkage disequilibrium score regression.

**RESULTS:** 97 phenotypes were significantly (false discovery rate < 0.05) and strongly (standardized effect size > |0.5|) associated with DSST performance in the discovery phase. Of those, 60 (38 with additional adjustment for education) demonstrated both statistical significance and consistent direction of association in the replication sample. The significantly associated phenotypes largely clustered into the following categories: psychiatric traits (e.g. anxiety, β per 1 SD = −0.74, P-value=3.9×10^−169^), education (e.g. highest math class completed, β per 1 SD = 2.11, P-value<1 × 10^−300^), leisure activities (e.g. solitary activities like puzzles, β per 1 SD = 1.85, P-value<1.0×10^−300^), social determinants (e.g. household income, β per 1 SD = 1.20, P-value= 8.9×10^−245^, and lifestyle (e.g. years smoked, β per 1 SD = −0.98, P-value= 2.2×10^−78^). We identified several reproducible genetic correlations between DSST and its top associated exposures (e.g. 0.48 for leisure activities like puzzles, 0.28 for years of education, and −0.24 for anxiety; all P ≤ 7.9×10^−26^). For almost all exposures, genetic correlations with DSST were considerably stronger than environmental correlations.

**CONCLUSIONS:** We have conducted the largest study of cognitive performance to date, building evidence supporting its correlations with many social, lifestyle, and clinical exposures. We established that the observed associations are in part underpinned by shared genetic architecture. Our study illustrates the potential of large-scale digital cohorts to contribute to epidemiologic discovery.

## Introduction

The advent of web-based data collection has dramatically transformed the landscape of epidemiologic studies, allowing fast and cost-effective phenotype ascertainment at an unprecedentedly large scale. The promise of this technology is well illustrated by behavioral, perceptual, and cognitive research. Even with potential noise introduced by device or hardware variability, digital assessments offer enhanced precision and greater accessibility at a lower cost (1). Consequently, the field of psychometrics has been steadily transitioning towards digital, specifically web-based, implementation of classic neurocognitive tasks.

The Digit Symbol Substitution Test (DSST), a widely used measure of cognitive processing speed, has been successfully computerized (2) and administered online (3). DSST asks a participant to match symbols to numbers according to a provided key; performance is operationalized as the number of correct matches over a predefined time interval (usually 90 seconds). A compelling body of evidence characterizes DSST as valid, reliable, brief, and especially sensitive to changes in cognitive function over time (4). In addition to capturing neurocognitive variability, DSST scores (adjusted for age and other demographic factors) have been prospectively linked to cardiovascular risk (5) and overall mortality (6). Performance on DSST is inversely correlated with age (7) and male sex (8), and has a substantial genetic component (twin study-derived h^2^=0.62) (9). However, evidence in support of other potential contributors, including education, medical conditions, or lifestyle factors, is either inconsistent across studies or does not yet exist. A deeper understanding of environmental contributions to DSST performance is essential for rigorous applications of this test in clinical and epidemiologic studies, e.g. for unbiased and precise estimation of associated disease risk, or for planning neurocognitive interventions.

To date, the 23andMe customer database contains the largest sample of DSST completions (N>200,000), coupled with a broad spectrum of genetic and environmental data. Leveraging this unique resource, we have conducted the first comprehensive, environment-wide study (ENWAS) of DSST performance in the 50-85 year age group. We integrated our findings with genotype data to estimate genetic correlations between DSST performance and its top environmental correlates, and compared genetic and environmental contributions to the observed phenotypic correlations. Finally, we evaluated reproducibility of our findings in an independent subsample of the 23andMe dataset. Our manuscript highlights the feasibility of administering DSST online in large-scale studies, as well as the utility of the 23andMe database to advance etiologic understanding of cognitive function.

## Methods

Throughout the study, we used R (10) for all statistical analyses and figures.

### Study population

The study population comprised 23andMe customers of European ancestry between 50 and 85 years of age, who resided in the United States, provided informed consent, and completed the DSST task online. Due to the visual nature of the DSST, we excluded participants who self-reported serious vision deficiencies, including age-related macular degeneration, retinal vein occlusion, and retinitis pigmentosa. Differences in demographic and clinical covariates were compared using t-tests for continuous variables and Chi-squared tests for categorical variables. The human subjects research protocol was approved by Ethical and Independent Review Services (www.eandireview.com), an external institutional review board accredited by the Association for the Accreditation of Human Research Protection Programs. This study complied with the principles outlined in the Declaration of Helsinki.

### Trait selection for the ENWAS

All traits tested for association with DSST were ascertained via web- or mobile-based surveys developed either in-house by 23andMe or previously published (e.g. the Pittsburgh Sleep Quality Index, PSQI)(11). We started with a broad range of phenotypes including health behaviors, personality traits, socioeconomic status, demographic variables, or clinical conditions. We subsequently included additional phenotypes derived from surveys focused on sleep, nutrition, and leisure activities. We excluded phenotypes that were later used as covariates in the ENWAS association models (e.g. age), traits with a predominantly genetic etiology (e.g. eye color), or those pertaining to medication response. These selection procedures yielded an initial list of 1033 traits. To avoid redundancy, we computed pairwise correlation coefficients for the 1033 initial phenotypes. For pairs that were highly correlated (|r|>0.80), we removed the phenotype with the smaller sample size to maximize statistical power. We further removed traits for which models would fail to converge due to lack of variation in the sample. The final phenotype list for the ENWAS included 824 traits.

### DSST administration and quality control

Two versions of DSST were deployed to our participants via the 23andMe website: one developed externally (the discovery dataset) and one developed in-house following identical specifications (the replication dataset). In both versions, the participants were presented with a key linking numbers and nine symbols (chosen at random from a symbol bank). They were first given the opportunity to practice selecting the correctly paired number for each of the four symbols sequentially shown on the screen. Following the practice round, the participants were asked to indicate as many correct pairings of numbers to symbols as possible during a 90 second interval.

The DSST score (i.e. number of correct trials per 90 seconds) distribution did not significantly deviate from normal, thus we did not transform the outcome. We excluded participants deemed outliers in DSST performance (100 or more correct trials per 90 seconds). After exclusions, we had data from a total of 238,214 individuals in the 50-85 age range who had completed the DSST game by the time of the analysis. Given the importance of age to cognitive outcomes and its potential for non-linear effects, we used loess regression to model the relationship between DSST performance and age in the total sample, stratified by sex.

### ENWAS

We modeled environment-wide associations using linear regressions with the number of correct DSST trials per 90 seconds as the outcome and each of the phenotypes as the main predictor, adjusted for age, sex, age*sex, device (desktop vs. laptop), five ancestry principal components, and version of the genotyping platform (used as a proxy for time of cohort entry). The model fit as a sensitivity analysis further adjusted for self-reported years of education. For the phenotypes that were sex-specific (e.g. age at first menses or azoospermia), the models above excluded sex and its interaction term with age. To protect against possible model misspecification, standard errors for all association estimates were derived from robust sandwich variance estimators. We additionally computed standardized (i.e. expressed per one standard deviation of the exposure) association estimates in each model to facilitate comparisons across traits. P-values were adjusted using the Benjamini-Yakutieli false discovery rate (FDR) approach to account for the residual phenotypic correlations, and adjusted P < 0.05 was used as a cut-off. We additionally filtered on post-stratified standardized effect size estimates, with |0.5| as a cut-off to ensure a meaningful association. Exposures that met these two criteria were carried forward to the replication phase. Models including identical covariates were fit to the discovery and replication data sets. Criteria for successful replication were set as 1) FDR P < 0.05 and 2) same direction of association in both discovery and replication analyses.

### Post-stratification of environment-wide association estimates

Data on the 824 putative correlates of DSST were collected via many different surveys, and most 23andMe participants typically only complete a subset of these surveys. Therefore, sample sizes for each environment-wide model varied widely. Moreover, the subsamples varied in their distributions of key demographic covariates: age, sex, and education. Because of these imbalances, we considered the total sample with available DSST data as the reference population and post-stratified association estimates for each environment-wide model. To create weights for post-stratification, we started with proportions of age groups (50-59, 60-69, and 70-85 years) and education (college-educated or not); unless the predictor was sex-specific (e.g. age at first menses), weights also included the male vs. female proportion. We then employed the iterative raking algorithm (12), which converges on a set of weights that ensure that the joint distribution of the post-stratification variables (i.e. age, sex, and education) matches that in the total DSST sample. The sandwich variance estimator described earlier was used to approximate the standard error of each post-stratified association estimate.

### Genetic correlations

We leveraged available genotype data on all participants (previously described in (13)) to explore the shared genetic architecture of cognitive performance and its top phenotypic correlates. We estimated genetic correlations between the DSST score and 32 phenotypes using linkage disequilibrium (LD) score regression (14), a widely used method predicated on the relationship between association test statistics for a given variant and its LD score under a polygenic model. Because most replicated correlates of DSST could be assigned to distinct groups (e.g. socioeconomic variables, health behaviors, psychiatric traits), we chose representative traits from each group for the genetic correlation analysis, taking into account sample size and strength of the association. We also included two negative controls, i.e. traits we found to be unassociated with DSST (red meat intake and kidney stones), for which we would expect a null genetic correlation. Advantages of the LD score regression approach include computational efficiency and validity under the condition of sample overlap, which was observed in our data. Genetic correlations were computed separately in the discovery and replication data sets; similarly to the environment-wide association study, successful replication was defined as P < multiple-testing corrected significance threshold and consistent direction of the genetic correlation across samples. The Bonferroni correction was applied to adjust for multiple comparisons.

### Environmental correlations

In addition to genetic correlations, we also estimated environmental correlations between DSST and its top environmental correlates using the formula proposed by Sodini *et al*.(15): 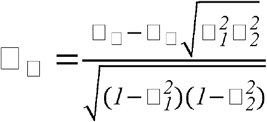, where □_□_ is the age- and sex-adjusted partial correlation coefficient between DSST and the traits of interest (same traits as in the genetic correlation analyses), □_□_ is genetic correlation as described above, and □*^2^*estimates refer to SNP-estimated heritabilities of DSST 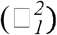 and the traits of interest 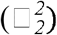. To explore the relative importance of genetic vs. environmental factors, we plotted □_□_ against □_□_ for the top traits. Like in the genetic correlation analysis, environmental correlations were estimated separately in the discovery and replication data sets.

## Results

### Study population

Although the discovery and replication subsamples came from the same source population of 23andMe customers, they were recruited during non-overlapping time periods: November 2016– November 2018 (discovery, n=144,786) and November 2018– July 2019 (replication, n=93,428). Relevant characteristics of both cohorts, stratified by sex, are presented in **Table 1**.

**Table 1:** Demographic and cognitive characteristics of the 23andMe research participants providing DSST data, stratified by sex. Continuous variables are presented as median [interquartile range], categorical variables as N (% total).

|  | Discovery (N=144,786) |  |  | Replication (N=93,428) |  |  |
| --- | --- | --- | --- | --- | --- | --- |
|  | Male | Female | N (% total) missing data | Male | Female | N (% total) missing data |
| N, % total | 64,415 (44.5%) | 80,371 (55.5%) | – | 39,626 (42.4%) | 53,802 (57.6%) | – |
| Age, years | 65 [58–71] | 63 [57–69] | – | 65 [58–71] | 63 [57–69] | – |
| Education, N (% total) |  |  | 6,349 (4.4%) |  |  | 17,594 (18.8%) |
| High school diploma or less | 4,076 (2.8%) | 7,557 (5.2%) |  | 2,549 (2.7%) | 5,143 (5.5%) |  |
| College diploma | 35,269 (24.4%) | 47,912 (33.1%) |  | 18,577 (19.9%) | 27,111 (29.0%) |  |
| Postgraduate degree | 22,346 (15.4%) | 21,277 (14.7%) |  | 11,496 (12.3%) | 10,958 (11.7%) |  |
| Cognitive game performance<br>DSST, # correct/90s | 47 [40–56] | 52 [43–58] | – | 44 [38–52] | 47 [40–54] | – |
| Device, N (% total) |  |  | 7,334 (5.1%) |  |  | 1,274 (1.4%) |
| Desktop | 33,874 (23.4%) | 36,805 (25.4%) |  | 21,970 (23.5%) | 25,713 (27.5%) |  |
| Laptop | 26,884 (18.6%) | 39,889 (27.6%) |  | 17,152 (18.4%) | 27,319 (29.2%) |  |
| Annual household income, USD |  |  | 68,074 (47.0%) |  |  | 68,876 (73.7%) |
| < 100,000 | 12,771 (8.8%) | 21,768 (15.0%) |  | 4,041 (4.3%) | 7,046 (7.5%) |  |
| >=100,000 | 21,788 (15.0%) | 20,385 (14.1%) |  | 7,168 (7.7%) | 6,297 (6.7%) |  |
| Body mass index, kg/m<sup>2</sup> | 28.2 (25.6 – 31.6) | 27.3 (23.6 – 32.0) | 3,804 (2.6%) | 28.2 (25.6 – 31.7) | 27.2 (23.5 – 31.7) | 2,973 (3.2%) |
| Ever tobacco user, % of total |  |  | 4,889 (3.4%) |  |  | 3,236 (3.5%) |
| No | 35,152 (24.3%) | 45,816 (31.6%) |  | 21,458 (23.0%) | 30,134 (32.3%) |  |
| Yes | 27,150 (18.8%) | 31,779 (21.9%) |  | 16,903 (18.1%) | 21,697 (23.2%) |  |

The majority of study participants were female, reflecting the sex breakdown of the source population of 23andMe customers. As DSST was targeted to individuals who were 50 years of age or older, median participant age was in the mid-60s. Compared to the overall US population in the 50+ year age group (16), our sample was approximately twice as likely to graduate from college. However, lifestyle characteristics (namely the percentage of never smokers and median BMI) were consistent with nationwide estimates (17, 18). Participants had the option to complete the task on either desktops or laptops; women were more likely to report the latter. Average performance on the DSST was comparable to that reported in a prior meta-analysis of data from 3,876 older adults (7). Similarly consistent with prior literature (8, 19), women on average performed better than men. In the combined discovery and replication sample, we observed a significant interaction between age and sex on DSST performance, with women exhibiting a slower age-related decline in average DSST performance (**Supplementary Figure 1**).

Most tests comparing demographics between discovery and replication cohorts were statistically significant; however, sample sizes of this magnitude typically lead to small P-values without meaningful differences. For example, the P-value comparing age distributions was 3.2×10^−15^, while the mean was 64.3 years in the discovery cohort and 64.0 in the replication cohort. Hence Table 1 does not contain P-values comparing the two subcohorts but rather shows measures of central tendency and dispersion for continuous variables and subgroup proportions for categorical variables. An important difference was observed for income and education, which were more likely to be missing in the replication cohort. This phenomenon arose from a change in data collection strategy: collecting data on socioeconomic variables was deprioritized.

### ENWAS

Definitions of all traits used in the ENWAS are provided in **Supplementary Text 1**. Subcohorts for each trait, which contained all participants who completed the DSST and had data on that specific trait as well as model covariates, ranged in size from 200 to 132,281 (discovery) and 948 to 74,744 (replication). We did not consider exposures for which N<200 for reasons of statistical power. Of the 824 exposures considered in the study, 557 were significantly (FDR < 0.05) associated with the DSST outcome in the discovery cohort after adjustment for relevant covariates (age, sex, proxy variable for cohort entry, age*sex interaction, and ancestry principal components) (**Supplementary Table 1a**). Additional adjustment for education (defined as less than college/college or above) attenuated most estimates of association and reduced the number of significant associations to 532. We post-stratified the trait-specific estimates to the overall DSST sample on age, sex, and education (only age and sex in education-adjusted models). Post-stratification did not meaningfully impact association estimates for most variables, except for those collected on smaller (N<5,000) subsets of participants. Similarly and predictably, the largest shifts in variance estimates due to the implementation of the sandwich estimator also occurred in the smallest subcohorts.

To enable comparisons of associations across models and facilitate interpretation, we multiplied post-standardized regression coefficients by the standard deviation of the exposure; such standardized effect estimates ranged from −1.02 to 2.11 (−0.93 to 1.84 in education-adjusted models). We focused all subsequent analyses on exposures for which the absolute value of the post-stratified association estimate exceeded 0.50 – an effect size per SD of exposure comparable to that of some antidepressants or benzodiazepines in prior studies of DSST (4). In the discovery phase, 97 traits met this criterion in addition to statistical significance (67 traits when additionally adjusted for education). Top traits associated with DSST were grouped into the following categories: education/socioeconomic status (e.g. vocabulary test score), musical ability (e.g. self-reported ability to sing back a note), mental health traits (e.g. the depression anxiety stress scale (DASS) anxiety subscore (20)), other health traits (e.g. self-rated general health), lifestyle (e.g. the PSQI score (11) as a measure of sleep), and other (e.g. tendency to eat before sleep). Post-stratified and standardized association estimates for representative traits from each category, all of which achieved reproducibility in the replication phase as defined below, are presented in **Figure 1**.

**Figure 1.**
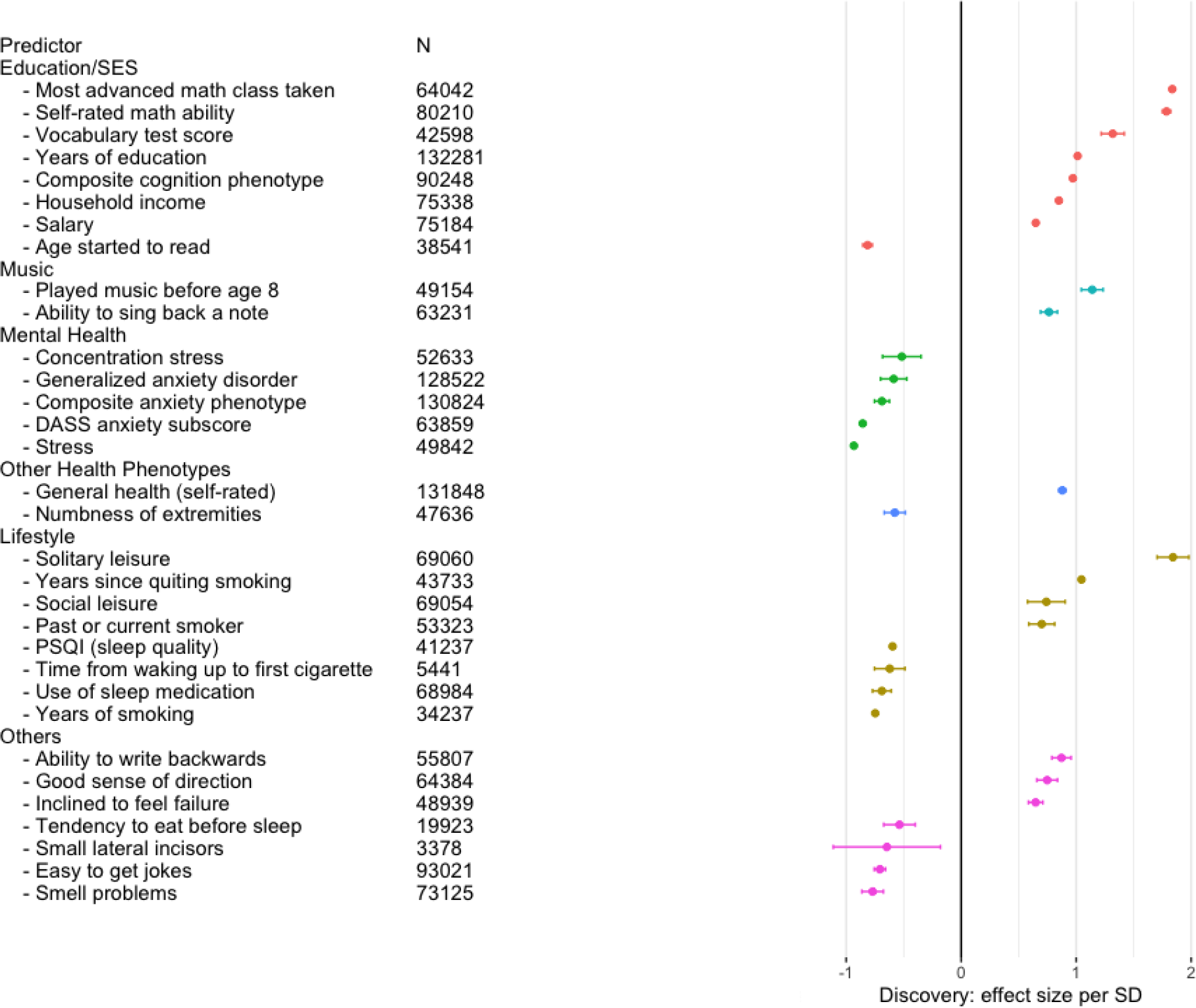
Top replicated associations between DSST performance and various exposures in the 23andMe cohort. Association estimates are grouped by etiologic theme, presented per one standard deviation of the independent variable, and shown with standard errors.

In the replication cohort, 95 traits (64 adjusted for education) achieved statistical significance (FDR < 0.05) and consistent direction of association, and 60 traits (38 adjusted for education) also exceeded the association size cutoff of |0.50| (**Supplementary Table 1b**).

Estimates of association for top predictors were highly concordant between discovery and replication studies, as shown for eight representative traits in **Figure 2**.

**Figure 2.**
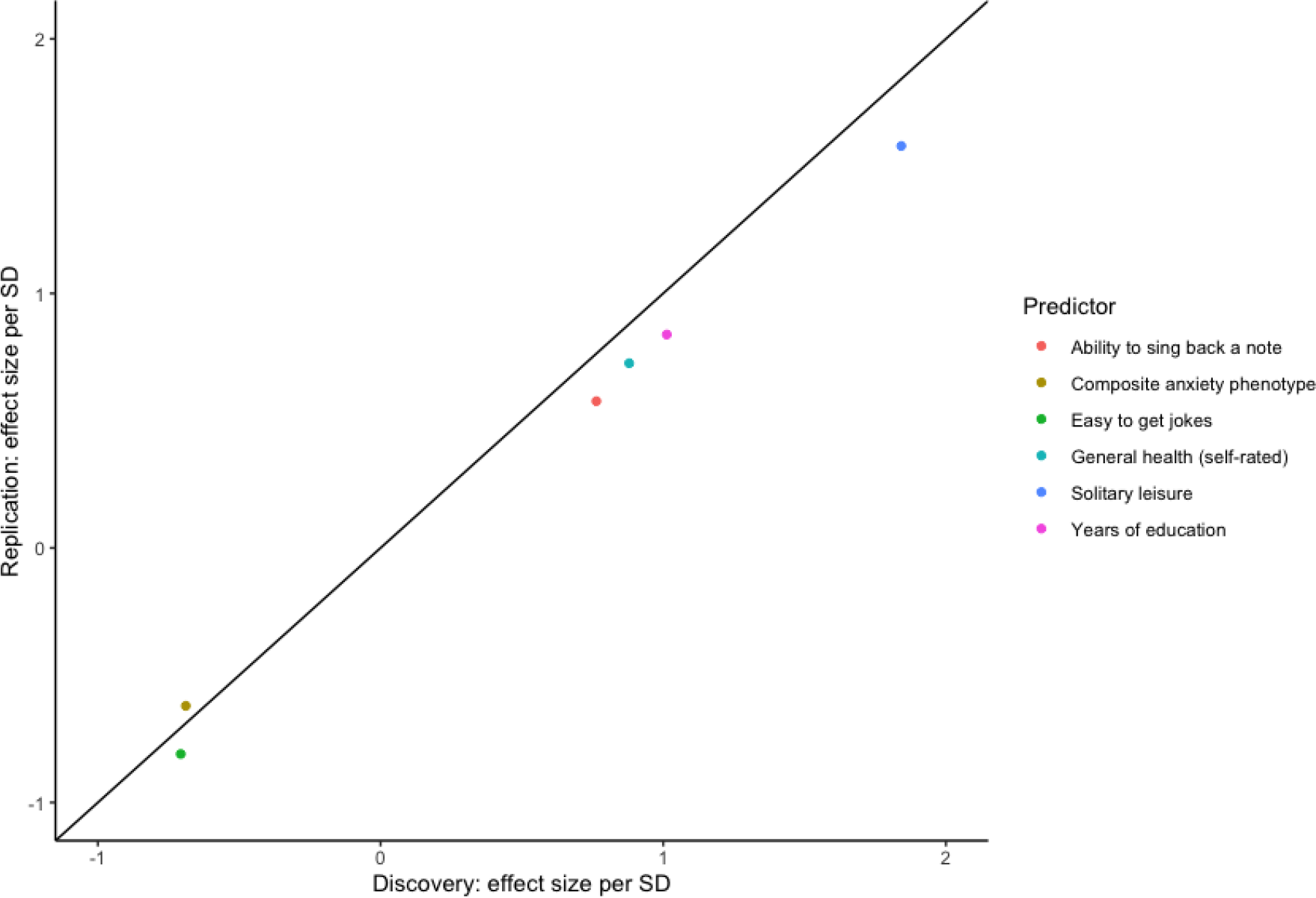
Concordance between standardized association estimates across the discovery and replication cohorts, presented for eight representative traits.

### Genetic and environmental correlations

Evidence for genetic and environmental underpinnings of the observed phenotypic associations for the traits shown in **Figure 1**, as well as for two negative controls (i.e. traits not found to be significantly associated with DSST and thus not expected to be genetically correlated with it, namely red meat intake and kidney stones), is presented in **Table 2** (discovery) and **Supplementary Table 2** (replication).

**Table 2:** Genetic and environmental correlations between top traits associated with DSST in the discovery cohort. Replicated correlations are shown in bold; criteria for replication were consistent direction + P-value < Bonferroni-corrected threshold in both discovery and replication cohorts. Negative controls are shown in grey.

| Trait | $h^2$ | SE($h^2$) | $r_g$ | SE($r_g$) | $r_g$ P-value | $r_e$ | SE($r_e$) | $r_e$ P-value |
| --- | --- | --- | --- | --- | --- | --- | --- | --- |
| Solitary leisure activities (e.g. puzzles) | 0.10 | 0.004 | **0.48** | **0.02** | **$7.4 \times 10^{-91}$** | **0.15** | **0.02** | **$1.7 \times 10^{-10}$** |
| Mathematical ability (self-rated) | 0.16 | 0.005 | **0.48** | **0.02** | **$5.4 \times 10^{-135}$** | **0.16** | **0.02** | **$7.5 \times 10^{-16}$** |
| Highest math course achieved | 0.17 | 0.005 | **0.45** | **0.02** | **$5.6 \times 10^{-111}$** | **0.17** | **0.02** | **$2.9 \times 10^{-17}$** |
| Played a musical instrument before age 8 | 0.04 | 0.002 | **0.29** | **0.03** | **$7.9 \times 10^{-18}$** | 0.13 | 0.03 | $7.6 \times 10^{-5}$ |
| Years of education | 0.13 | 0.004 | **0.28** | **0.02** | **$5.5 \times 10^{-41}$** | **0.15** | **0.02** | **$8.8 \times 10^{-13}$** |
| Vocabulary test score | 0.10 | 0.004 | **0.28** | **0.03** | **$3.7 \times 10^{-23}$** | **0.15** | **0.03** | **$2.8 \times 10^{-8}$** |
| Cognition score | 0.07 | 0.004 | **0.27** | **0.03** | **$2.7 \times 10^{-17}$** | **0.10** | **0.03** | **0.001** |
| Years since quitting smoking | 0.06 | 0.003 | **0.26** | **0.03** | **$5.7 \times 10^{-18}$** | **0.11** | **0.03** | **0.0002** |
| Social leisure activities (e.g. card games with friends) | 0.04 | 0.003 | **0.23** | **0.04** | **$2.6 \times 10^{-10}$** | 0.06 | 0.04 | 0.09 |
| Ability to write backwards | 0.09 | 0.004 | **0.21** | **0.03** | **$4. \times 10^{-13}$** | 0.09 | 0.03 | 0.003 |
| Stopped smoking | 0.04 | 0.002 | **0.18** | **0.03** | **$1.3 \times 10^{-9}$** | 0.08 | 0.03 | 0.004 |
| Feel like a failure | 0.05 | 0.002 | 0.16 | 0.03 | $4.0 \times 10^{-8}$ | 0.07 | 0.03 | 0.01 |
| Sense of direction | 0.10 | 0.004 | **0.13** | **0.02** | **$2.1 \times 10^{-7}$** | 0.07 | 0.02 | 0.007 |
| General health | 0.09 | 0.003 | 0.12 | 0.02 | $1.6 \times 10^{-7}$ | **0.12** | **0.02** | **$5.9 \times 10^{-7}$** |
| Personal earnings during the past 12 months | 0.06 | 0.002 | 0.12 | 0.03 | $1.7 \times 10^{-5}$ | **0.10** | **0.03** | **0.0001** |
| Total household income | 0.07 | 0.003 | 0.11 | 0.03 | 0.003 | **0.13** | **0.03** | **$1.1 \times 10^{-6}$** |
| Ability to sing back musical notes | 0.16 | 0.005 | 0.07 | 0.02 | 0.002 | **0.10** | **0.02** | **$5.1 \times 10^{-5}$** |
| Red meat servings | 0.04 | 0.001 | 0.02 | 0.02 | 0.39 | -0.01 | 0.02 | 0.78 |
| Kidney stones | 0.05 | 0.003 | -0.04 | 0.03 | 0.08 | 0.00 | 0.03 | 0.99 |
| Ability to understand jokes | 0.06 | 0.003 | **-0.13** | **0.03** | **$3.0 \times 10^{-6}$** | -0.08 | 0.03 | 0.006 |
| Eating before bed | 0.13 | 0.006 | **-0.14** | **0.04** | **$7.1 \times 10^{-5}$** | -0.06 | 0.04 | 0.12 |
| Numbness in extremities | 0.05 | 0.002 | -0.16 | 0.03 | $6.6 \times 10^{-8}$ | -0.07 | 0.03 | 0.19 |
| First cigarette after waking | 0.08 | 0.004 | -0.18 | 0.03 | $1.0 \times 10^{-7}$ | -0.10 | 0.03 | 0.004 |
| Perceived stress | 0.09 | 0.004 | **-0.18** | **0.03** | **$2.7 \times 10^{-8}$** | -0.06 | 0.03 | 0.06 |
| Overall sleep quality | 0.10 | 0.014 | **-0.18** | **0.05** | **0.0001** | -0.08 | 0.05 | 0.08 |
| Depression Anxiety Stress Scales (DASS) - anxiety subscale | 0.06 | 0.002 | **-0.19** | **0.03** | **$1.5 \times 10^{-9}$** | -0.09 | 0.03 | 0.002 |
| Patient Health Questionnaire (PHQ) - concentration difficulties | 0.05 | 0.002 | **-0.19** | **0.03** | **$2.8 \times 10^{-8}$** | -0.05 | 0.03 | 0.11 |
| Anxiety | 0.06 | 0.002 | **-0.24** | **0.02** | **$7.9 \times 10^{-26}$** | -0.06 | 0.02 | 0.01 |
| Generalized anxiety disorder | 0.04 | 0.001 | **-0.25** | **0.03** | **$2.7 \times 10^{-21}$** | -0.05 | 0.03 | 0.07 |
| Years a smoker | 0.04 | 0.003 | **-0.25** | **0.04** | **$2.2 \times 10^{-10}$** | -0.09 | 0.04 | 0.03 |
| Age started reading | 0.04 | 0.002 | **-0.28** | **0.03** | **$6.6 \times 10^{-24}$** | -0.06 | 0.03 | 0.04 |
| Sleep medication usage | 0.08 | 0.004 | **-0.28** | **0.03** | **$4.8 \times 10^{-22}$** | -0.07 | 0.03 | 0.02 |
| Impaired smell function | 0.03 | 0.003 | **-0.29** | **0.04** | **$1.8 \times 10^{-12}$** | -0.08 | 0.04 | 0.05 |

In the LD score regression analyses, heritability of DSST performance was estimated at 0.16 (SE=0.008). LD score regression performs less reliably for traits with lower heritability; therefore, Bulik-Sullivan et al. (14) have proposed a heritability Z-score cutoff of 4. One variable in our data set (small lateral incisors) had a Z-score < 4 and was removed; therefore, 31 traits were retained for the correlation analysis. Exposure heritabilities varied from 0.03 (self-reported sleep problems) to 0.17 (highest math class taken). As expected, DSST was neither genetically nor environmentally correlated with the negative controls. Overall, 30 traits had a significant genetic correlation and 12 had a significant environmental correlation with DSST; of those, 24 genetic correlations and 11 environmental correlations were successfully replicated. As a general pattern with few exceptions (e.g. household income), estimates of genetic correlation exceeded the estimates of environmental correlation, in some cases by an order of magnitude (**Figure 3**).

**Figure 3.**
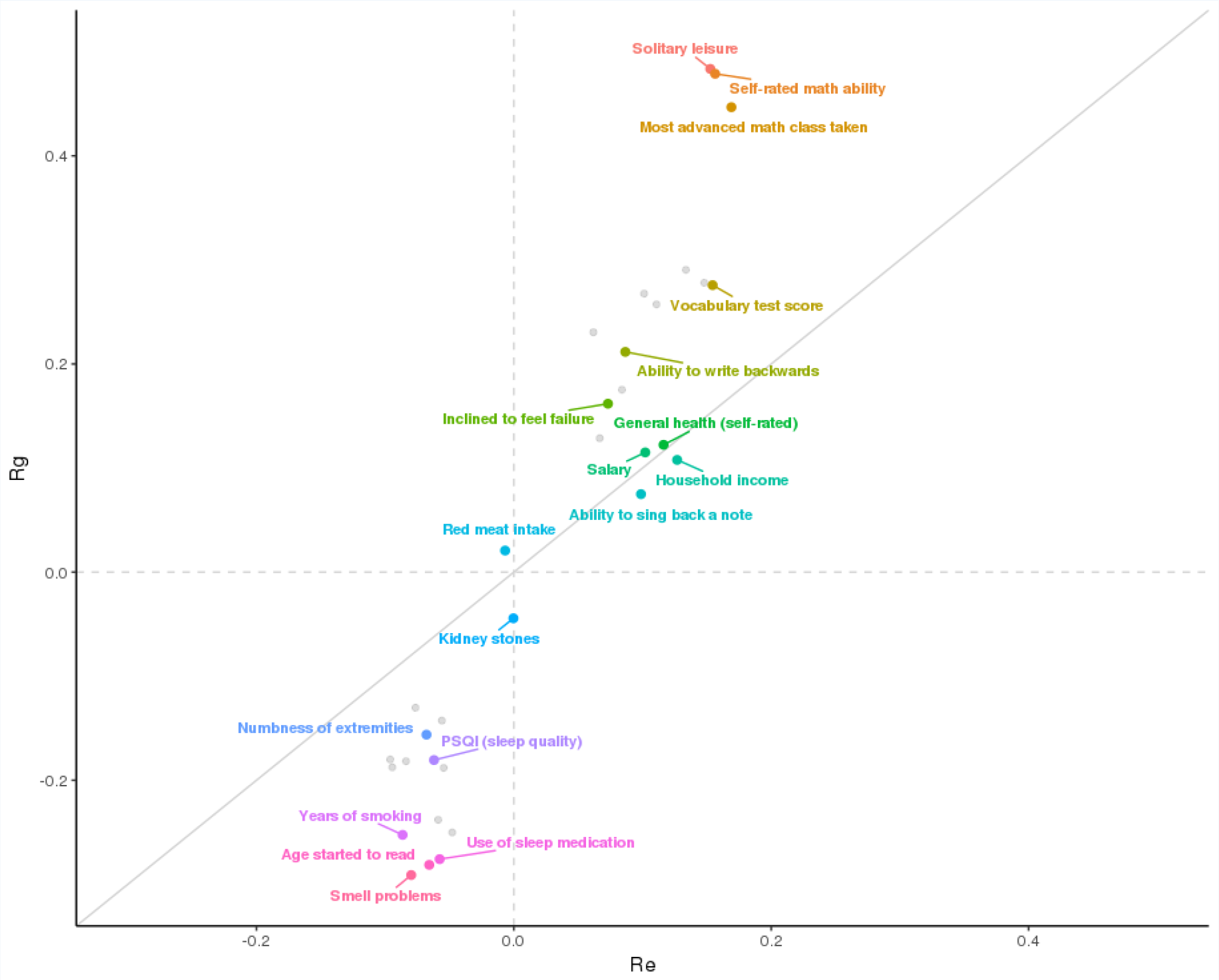
Genetic (r_g_) vs. environmental (r_e_) correlations for 31 top replicated traits with heritability Z-scores > 4. Each data point corresponds to one trait; some representative traits are shown in color.

## Discussion

Using the ENWAS framework, we carried out the first comprehensive investigation of non-genetic contributions to cognitive performance in a large sample of US-based 23andMe research customers of European descent. We identified numerous reproducible associations between DSST scores and predictors that are both known (e.g. smoking (21) or sleep (22)) and novel (e.g. mathematical ability or numbness of extremities). Leveraging genotype data available on the same individuals, we also showed that the observed phenotypic correlations are in part due to the shared genetic architecture of DSST performance and its top environmental predictors.

We chose DSST as a metric of cognitive performance for several reasons. DSST is easily administered via the web, and reliable (23); it is indicative of multiple cognitive functions, including processing speed, attention, visual and spatial perception, and executive function (24). Optimally for our cohort, DSST offers higher sensitivity at moderate to high levels of cognition compared to other tests, e.g. the Mini Mental State Examination (MMSE) (25). Notably, DSST performance is widely believed to be only minimally affected by linguistic, cultural, or educational factors (4). However, most associations observed in our study were sensitive to adjustment for educational achievement, and measures of socioeconomic status and education prominently figured on the list of top associations, indicating that the impact of education may not be so minimal after all. The underlying mechanisms likely include both direct effects of education on cognition and cognitive effects of socioeconomic status, for which education may be used as a proxy. For example, the latter (socioeconomic) effect may be mediated by stress, triggered by financial insecurity and known to affect DSST performance (26). Consistent with this hypothesis, perceived stress was strongly (standardized effect size: −1.02, P=2.30*10^−120^) and reproducibly associated with DSST performance in our sample. As for the former (direct) effect, education has previously been postulated to increase cognitive reserve, defined as an individual’s ability to maintain higher function despite onset of various brain pathologies (27), by increasing controlled (i.e. not automatic) cognitive processes and conceptualization abilities (28). Although the buffering role of cognitive reserve has been established for most chronic brain pathologies (27), previous investigations of the effect of education on DSST performance yielded mixed results. In some disease settings (e.g. Alzheimer’s (29)), evidence favors a protective effect of education on age-related decline in DSST performance, and in others (e.g. brain infarct (30)) such effects are null. In contrast to disease-focused studies, our sample was recruited from the general population of 50-85 year old 23andMe customers rather than a neurodegenerative disease cohort, enriched for college-educated individuals, and restricted to individuals who could independently complete at least one survey and the DSST task. Consistently with our observations, education was strongly associated with DSST performance in other healthy populations (8). Our findings highlight the importance of education and socioeconomic status to DSST performance in non-clinical settings, and suggest that future studies of DSST would benefit from accounting for educational heterogeneity in both design and analysis.

The ENWAS paradigm provides a useful way to systematically and agnostically interrogate many potential associations while properly controlling for multiple testing. It is especially well suited for large-scale cohorts with a broad spectrum of phenotypic measurements, such as the 23andMe population. Several methodological caveats, however, complicate the interpretation of such studies. A recent review by Manrai et al. (31) identified several threats to the validity of ENWAS findings: unadjusted confounding, challenges of measuring dynamic environmental exposures, the correlated nature of the exposures, and ‘excessive’ statistical power leading to detection of etiologically insignificant, small effects. We would like to add the difficulty of comparing effects across both traits and subpopulations to this list and offer several suggestions for ENWAS best practice. First, we ensured that our discovery results (both in ENWAS and the genetic/environmental correlation analyses) are reproducible in an *a priori* selected data set of independent individuals representing the same source population of 23andMe customers. Second, we standardized our association estimates to enable easy comparisons and prioritization of findings. Third, because the subsets of participants with information on each trait often varied in age, sex, and education, we post-stratified our association estimates on these variables to reduce the spurious effects of such unbalances. In our study, post-stratification and the use of robust variance estimators did not dramatically change our estimates for most subcohorts in our study because of their large sample size. Nevertheless, for subcohorts ranging from several hundred to several thousand (N<5,000) individuals (i.e. cohorts comparable in size to most epidemiologic studies), these procedures did have a pronounced effect and should be implemented to reduce bias and appropriately quantify uncertainty. Fourth, we filtered our findings on absolute standardized effect size, so that the impact of our reported findings exceeds that observed in experimental studies of clinically meaningful exposures (namely antidepressant use and sleep deprivation) on DSST performance. Such filtering reduces the likelihood that our findings were falsely positive (heeding Hill’s strength of association guideline (32)) and focuses the subsequent analyses on exposures that are likely to matter most for cognition. Fifth, we incorporated genomic data to probe some of the mechanisms underlying the observed associations, which can be challenging to do in a large-scale, cross-sectional study.

Specifically, we established that many of the replicated ENWAS associations are at least partially underpinned by shared genetic architecture. Estimates of genetic correlation were especially striking for engagement in solitary leisure activities like puzzles (r_g_=0.48, P=7.44×10^−91^), as well as for various metrics of mathematical and musical education (e.g. whether someone played music before the age of eight, r_g_=0.29, P=7.91×10^−18^). These are predictors that are commonly considered environmental rather than genetic. For example, exposure to music lessons in childhood is more likely to be construed as a marker of parental socioeconomic status rather than of innate ability. But even parental socioeconomic status is not an exclusively environmental predictor, because it is strongly correlated with parental educational attainment, which in turn has a genetic component (33) that is also passed down to the offspring. Therefore, it is not surprising to see statistically significant genetic correlations between DSST performance and measures of education/socioeconomic status. Crucially, numerous prior studies have demonstrated that genetic influences on cognition are more pronounced in a) older adults and b) more affluent populations (34): both conditions accurately characterize the 23andMe DSST cohort. Supporting this hypothesis, our estimates of environmental correlations between DSST and our variables of interest (e.g. for playing music before the age of eight, r_e_= 0.13, P= P=7.61×10^−5^) were significant yet consistently lower than their genetic counterparts. These findings may not generalize to cohorts with greater socioeconomic and educational diversity, where environmental factors account for a larger percentage of cognitive variability (35).

Combining the ENWAS framework with genomic data, we have identified multiple strong associations with DSST performance. Many of these associations have previously been reported; for example, the inverse relationship between psychiatric symptoms and DSST scores is well known. However, evidence supporting relationships between DSST performance and other health conditions or lifestyle variables is only beginning to emerge. A recent analysis of the UK Biobank data (36) has linked self-reported chronic inflammatory conditions, defined broadly to include musculoskeletal, endocrine, autoimmune, and other conditions, to decreased DSST performance. In our data set, self-reported administration of anti-inflammatory medication was similarly associated with lower DSST scores (standardized effect: −0.35, P-value= 4×10^−16^ in the discovery cohort), although it did not pass the effect size threshold we used to prioritize the associations. Moreover, our data set offers greater granularity of associations: among chronic conditions included under the UK Biobank’s umbrella definition of inflammatory, we note that the greatest reductions in DSST were observed for participants with type 2 diabetes, followed by COPD and rheumatoid arthritis (**Supplementary Table 1a**). Furthermore, we showed the associations with inflammatory conditions and DSST were considerably (>10%) attenuated upon adjustment for education attainment. The breadth of our database also enabled identifying novel inverse associations between chronic disease and cognitive performance, e.g. among patients with nephritis or back pain. Other novel and reproducible findings included positive relationships between mathematical and musical education earlier in life and DSST performance in older adulthood, as well as between current engagement with leisure activities (e.g. puzzles or computer games) and cognitive performance.

Because we are reporting on cross-sectional data, we are limited in inferring both temporality and causality from observed associations. For exposures pertaining to childhood or early adulthood (music lessons, education), the temporality of exposure preceding the DSST outcome is implied, because all of our participants were 50 years of age or older at the time of DSST. For many other exposures, cognitive ability (as measured by DSST) may affect both the quality of collected information (thus leading to bias) or be upstream of the exposures considered in our study (reverse causality). Another potential limitation is the possibility of residual confounding. Although we adjusted for education as the best proxy of socioeconomic status available on most individuals, the role of other potential confounders should be explored in future, more narrowly focused studies. Finally, the demographic composition of our cohort--mostly college-educated older individuals of European descent--limits generalizations to other, more diverse populations.

Like in previous ENWAS, the primary application of our findings lies in generating etiologic hypotheses and prioritizing non-genetic exposures for future studies in smaller, well phenotyped cohorts. In agreement with previous twin and SNP-based heritability studies, our analyses also show that many of the exposures identified in our study have a genetic component that can be leveraged in pursuit of greater etiologic understanding. For example, a recent Mendelian randomization study (37) used genetic anchors to investigate the causal relationship between sleep duration and cognition in the UK Biobank and found evidence of a non-linear relationship. These insights can inform interventions to improve cognition in older adults, focusing on modifiable factors with the strongest causal effects. However, the cost and effort involved in such interventions requires rigorous selection of appropriate exposures, and we demonstrated that ENWAS on large-scale cohorts can be its fruitful first step. In conclusion, the findings of our study showcase the utility of 1) the combined ENWAS + genomics approach and the 23andMe database to comprehensively interrogate contributions to cognitive performance in older adulthood.

## Data Availability

N/A

## Acknowledgements

We thank the 23andMe research participants who enabled this study, and Dr. Laura Germine for her assistance in developing the web-based cognitive test.

## Author Contributions

Y.H. conducted most of the data analyses. T.F. curated and summarized phenotype definitions, and created the supplementary tables. R.G. guided the selection of statistical methods and helped interpret results. S.A. designed and supervised the study, and wrote the manuscript. All authors reviewed the content of the manuscript. The 23andMe Research Team provided administrative, technical, or material support.

Members of the 23andMe Research Team include: Michelle Agee, Adam Auton, Robert K. Bell, Katarzyna Bryc, Sarah K. Clark, Sarah L. Elson, Kipper Fletez-Brant, Pierre Fontanillas, Nicholas A. Furlotte, Pooja M. Gandhi, Jessica Inchauspé, Naomi G. Iwata, Karl Heilbron, Barry Hicks, David A. Hinds, Karen E. Huber, Ethan M. Jewett, Yunxuan Jiang, Aaron Kleinman, Keng-Han Lin, Nadia K. Litterman, Karl Bo Lopker, Jennifer C. McCreight, Matthew H. McIntyre, Kimberly F. McManus, Joanna L. Mountain, Sahar V. Mozaffari, Priyanka Nandakumar, Elizabeth S. Noblin, Carrie A.M. Northover, Jared O’Connell, Steven J. Pitts, G. David Poznik, J. Fah Sathirapongsasuti, Anjali J. Shastri, Janie F. Shelton, Suyash Shringarpure, Chao Tian, Joyce Y. Tung, Robert J. Tunney, Vladimir Vacic, Xin Wang, Amir S. Zare.

---

Supplementary Figure 1. Loess curves illustrating DSST performance in the 23andMe sample by age and sex. DSST scores showed a steady decrease across age groups, with women exhibiting a slower decline after mid-50s.

